# Evaluation of the Multi-Cancer Early Detection (MCED) value of YiDiXie^™^-HS and YiDiXie^™^-SS

**DOI:** 10.1101/2024.03.11.24303683

**Authors:** Chen Sun, Chong Lu, Yongjian Zhang, Ling Wang, Zhenjian Ge, Zhenyu Wen, Wenkang Chen, Yingqi Li, Yutong Wu, Shengjie Lin, Pengwu Zhang, Wuping Wang, Siwei Chen, Huimei Zhou, Xutai Li, Shaobin Wang, Yong Xia, Wei Li, Wei Lan, Yongjiang Cai, Ling Ji, Haibo Wang, Yongqing Lai

## Abstract

Cancer is a serious threat to the whole of humanity. The Multi-Cancer Early Detection (MCED) test is expected to solve the problem of “Universal cancer screening”. The purpose of this study is to evaluate the MCED value of two MCED tests, YiDiXie™-HS and YiDiXie™-SS, in multiple cancer types.

**Patients and methods:** 11094 subjects were finally included in this study (the malignant tumor group, n = 4405; the normal control group, n = 6689). The malignant tumor group included all major solid and hematological malignant tumor types. The sensitivity and specificity of YiDiXie™-HS and YiDiXie™-SS were evaluated, respectively.

**Results:** The overall sensitivity of YiDiXie™-HS for different cancer types and stages was 90.1% (89.2% - 90.9%; 3971/4405), and its specificity was 89.7% (89.0% - 90.4%; 6002/6689). Its sensitivity increases with clinical stage: stage I, 85.6% (83.9% - 87.1%); stage II, 91.4% (89.6% - 93.0%); stage III, 93.9% (92.0% - 95.4%); and stage IV, 98.4% (96.9% - 99.2%). The overall sensitivity of YiDiXie™-SS for different cancer types and stages was 99.1% (98.8% - 99.3%; 4365/4405), and its specificity was 65.2% (64.0% - 66.3%; 4358/6689). Its sensitivity was basically comparable in each clinical stage: stage I, 98.6% (98.0% - 99.1%); stage II, 99.5% (98.9% - 99.8%); stage III, 99.5% (98.6% - 99.8%); stage IV, 99.8% (98.9% - 100.0%).

**Conclusion:** YiDiXie™-HS has a high sensitivity in all clinical stages of all cancer types. YiDiXie™-SS has an extremely high sensitivity in all clinical stages of all cancer types. YiDiXie™-HS and YiDiXie™-SS can replace existing cancer screening tests and are expected to solve the world problem of “Universal cancer screening”.

**Clinical trial number:** ChiCTR2200066840.

## INTRODUCTION

The World Health Organization’s International Agency for Research on Cancer (IARC) released the latest data on the global burden of cancer, showing that in 2022, there were 19.96 million new cancer cases and 9.70 million cancer deaths globally; there were 4.82 million new cancer cases and 2.57 million cancer deaths in China, with the number of new cancer cases and deaths ranking first in the world^1^. Therefore, cancer is a serious threat to Chinese and the whole of humanity.

Cancer imposes a heavy financial burden on society, with total cancer treatment expenditures in the United States alone exceeding $200 billion in 2020^2-4^, and projected to reach $246 billion in 2030^5^. In addition, cancer treatment imposes a significant financial burden on patients, with patient out-of-pocket expenditures for cancer treatment in the United States estimated at $16 billion annually^6^; 12% to 62% of cancer patients in the United States are reported being in debt because of their treatment^7^. Thus, cancer places a heavy financial burden on society and patients.

“Universal cancer screening” can significantly improve patient prognosis^8-14^, dramatically reduces socio-economic burdens^15-17^, markedly improves patients’ economic status^6,16,18^ and significantly improve patient employment^19^.

However, the screening model of existing cancer screening tests does not fulfill the need for “Universal cancer screening”. This screening model^20^ can be referred to as the model of “Single-Cancer Early Detection (SCED)”, which refers to the application of existing cancer screening tests (e.g., CT, ultrasound, gastroscopy, colonoscopy, blood TPSA, etc.) to screen for one site-specific cancer at a time (e.g., CT scan for lung cancer, mammogram for breast cancer, TPSA for blood, etc.), and Subjects undergo multiple examinations or tests to screen for multiple cancers.

Several shortcomings of the SCED model limit its use in “Universal cancer screening”. First, the public often forgoes cancer screening due to concerns that the screening process is too cumbersome^16^, the tests are expensive^21^, and some of the tests are invasive and radioactive^21-25^. Second, the SCED model does not enable “Universal cancer screening” due to the low incidence of the individual cancer types screened. Conventional guidelines recommend “high-incidence cancer screening for high-risk populations” not “universal cancer screening”^26-30^, which consequently leads to a poorer prognosis for the majority of cancer cases^8,9,12,31^. Finally, the SCED model leads to a large accumulation of false-positive results^32^ and significantly increases patient anxiety^33,34^ and subsequent medical costs^32,35^. Therefore, there is an urgent need to find a better screening model to fulfill the need of “Universal cancer screening”.

Recently, a new blood test called the “Multi-Cancer Early Detection (MCED) test” or “Pan-cancer test” has been developed, which allows for the early detection of multiple cancers with a single blood test ^36-38^. They typically combine artificial intelligence (AI) and machine learning with the detection of a variety of circulating analytes, including free cell DNA (cfDNA), circulating tumor cells (CTCs), miRNAs, exosomes, and others, to detect early signs of multiple cancers^37-39^. These MCED tests have generated a new model of cancer screening, the MCED model, and is expected to solve the world’s problem of “Universal cancer screening”.

Based on the detection of miRNAs in serum, Shenzhen KeRuiDa Health Technology Co., Ltd. has developed “YiDiXie ™ all-cancer test” (hereinafter referred to as the “YiDiXie™ test”). With only 200 milliliters of whole blood or 100 milliliters of serum, the test can detect multiple cancer types, enabling early detection of cancer at home. The “YiDiXie ™ test” consists of three independent tests: YiDiXie™ -HS, YiDiXie™-SS and YiDiXie™-D.

The purpose of this study is to evaluate the MCED value of two MCED tests, YiDiXie™-HS and YiDiXie™-SS in multiple cancer types.

## PATIENTS AND METHODS

### Study design

The SZ-PILOT study (ChiCTR2200066840) was a single-center, prospective, observational study. Subjects who signed the broad informed consent for donation of remaining samples at the time of admission or medical health checkup were included, and 0.5 ml of their remaining serum samples were collected for this study.

This study was blinded. Neither the laboratory personnel performing the “YiDiXie™ test” nor the technicians of KeRuiDa Co. evaluating the raw results of the “YiDiXie™ test” were informed of the subject’s clinical information. The clinical experts assessing the subjects’ clinical information were also unaware of the results of the “YiDiXie™ test”.

The study was approved by the Ethics Committee of Peking University Shenzhen Hospital and was conducted in accordance with the International Conference on Harmonization for “Good clinical practice guidelines” and the Declaration of Helsinki.

### Participants

Subjects in the two groups were enrolled separately, and all subjects who met the inclusion criteria were included consecutively.

The malignant tumor group initially enrolled hospitalized patients with “suspected (solid or hematological) malignant tumors” with a signed broad informed consent for donation of the remaining samples. Subjects with tumor that has been surgically removed or disappeared after treatment at the time of sample acquisition, no surgical or biopsy pathology diagnosis, or ambiguous pathology were excluded from the malignant tumor group.

The normal control group initially included healthy medical examiners signing a broad informed consent for donation of the remaining samples. Subjects with undiagnosed suspected malignant tumors were excluded from the normal control group.

Subjects who were not qualified in the serum sample quality test prior to the “YiDiXie ™ test” were excluded from the study.

### Sample collection, processing

The serum samples used in this study were obtained from serum left over after a normal consultation, without the need for additional blood sampling. Approximately 0.5 ml of serum was collected from the remaining serum of the participants in the Medical Laboratory and stored at - 80°C for use in the subsequent “YiDiXie™ test”.

### “YiDiXie™ test”

The “YiDiXie ™ test” is performed using the “YiDiXie™ all-cancer detection kit”. The “YiDiXie™ all-cancer detection kit” is an in-vitro diagnostic kit developed and manufactured by Shenzhen KeRuiDa Health Technology Co., Ltd. for use in fluorescent quantitative PCR instruments. It detects the expression levels of dozens of miRNA biomarkers in serum to determine whether cancer is present in the subject. It predefines appropriate thresholds for each miRNA biomarker, ensuring that each miRNA marker has a high specificity ( ≥ 0.95). The YiDiXie ™ kit integrates these independent assays in a concurrent testing model to significantly increase the sensitivity in broad-spectrum cancers and maintain a high specificity.

The “YiDiXie™ test” consists of three tests with highly different characteristics: YiDiXie ™ -HS, YiDiXie ™ -SS and YiDiXie ™ -D. The YiDiXie ™ -HS (YiDiXie™-Highly Sensitive) is the standard version of the “YiDiXie™ test”, which was developed with high sensitivity and high specificity. On the basis of YiDiXie ™ -HS, YiDiXie ™ -SS (YiDiXie ™ -Super Sensitive) significantly increases the number of miRNA tests to achieve extremely high sensitivity for all stages in all malignancy types. Based on YiDiXie ™ -HS, YiDiXie ™ -D (YiDiXie ™ -Diagnosis) significantly increases the diagnostic threshold of individual miRNA tests to achieve very high tumors, and therefore its early cancer screening performance was not evaluated in this study.

Perform the “YiDiXie™ test” according to the instructions of the “YiDiXie™ all-cancer detection kit”. Briefly, take 20 μ l of serum, add 20 μ l of Nucleic Acid Extract, mix well and centrifuge at 50 ° C for 20 minutes, 95 °C for 5 minutes, and 13,000 rpm at 4 °C for 5 minutes, and the supernatant is the Nucleic Acid Extract. Take 8 μl of crude nucleic acid extract, add 12 μ l of reverse transcription reaction solution, mix well, keep warm at 37 °C for 30 min, keep warm at 42 ° C for 30 min, heat at specificity. YiDiXie™-D is designed for preoperative diagnosis of a wide range of 75 °C for 5 min, and leave on ice for 2 min. cDNA was diluted by 20-fold for further analysis. Take 4 μ l of cDNA dilution solution, add 6 μ l of amplification solution, mix well, and then carry out RT-qPCR reaction program. The RT-qPCR running program was set up as follows: 95 ° C for 2 min, then 40 cycles of 95 °C for 10s, 60 °C for 30 s and 70 °C for 30s.

The original test results were analyzed by the laboratory technicians of KeRuiDa Co. and determined to be “positive” or “negative”.

### Clinical data collection

Clinical, pathological, laboratory, and imaging data in this study were extracted from the subjects’ hospitalized medical records or physical examination reports. Clinical staging was completed by trained clinicians assessed according to the AJCC staging manual (seventh or eighth edition) ^40,41^.

### Statistical analyses

For demographic and baseline characteristics, descriptive statistics were reported. For categorical variables, the number and percentage of participants in each category were calculated; for continuous variables, the total number of participants (n), mean, standard deviation (SD) or standard error (SE), median, first quartile (Q1), third quartile (Q3), minimum, and maximum values were calculated. The 95% confidence intervals (CIs) for multiple indicators were calculated using the Wilson (score) method.

## RESULTS

### Participant disposition

A total of 11754 study subjects (The malignant tumor group, n = 4963; The normal control group, n = 6791) were initially enrolled in this study (Fig. 1). 102 cases in the normal control group were excluded due to undiagnosed suspected tumors. A total of 558 cases were excluded from the malignant tumor group, of which 345 cases had no pathological results, 170 cases with tumors surgically removed or regressed after treatment, and 43 cases had ambiguous benign or malignant pathological results. There were no samples that failed the test due to substandard serum quality, which was mainly because the samples used in this study were residual serum after regular test, substandard samples had been excluded by the medical laboratory, and the samples were stored under good conditions. Mild hemolysis or samples stored at unsuitable temperatures can lead to test failure. All exclusion categories were preset before enrollment. This study finally included 11094 study subjects (The malignant tumor group, n = 4405; The normal control group, n = 6689).

**Figure 1.**
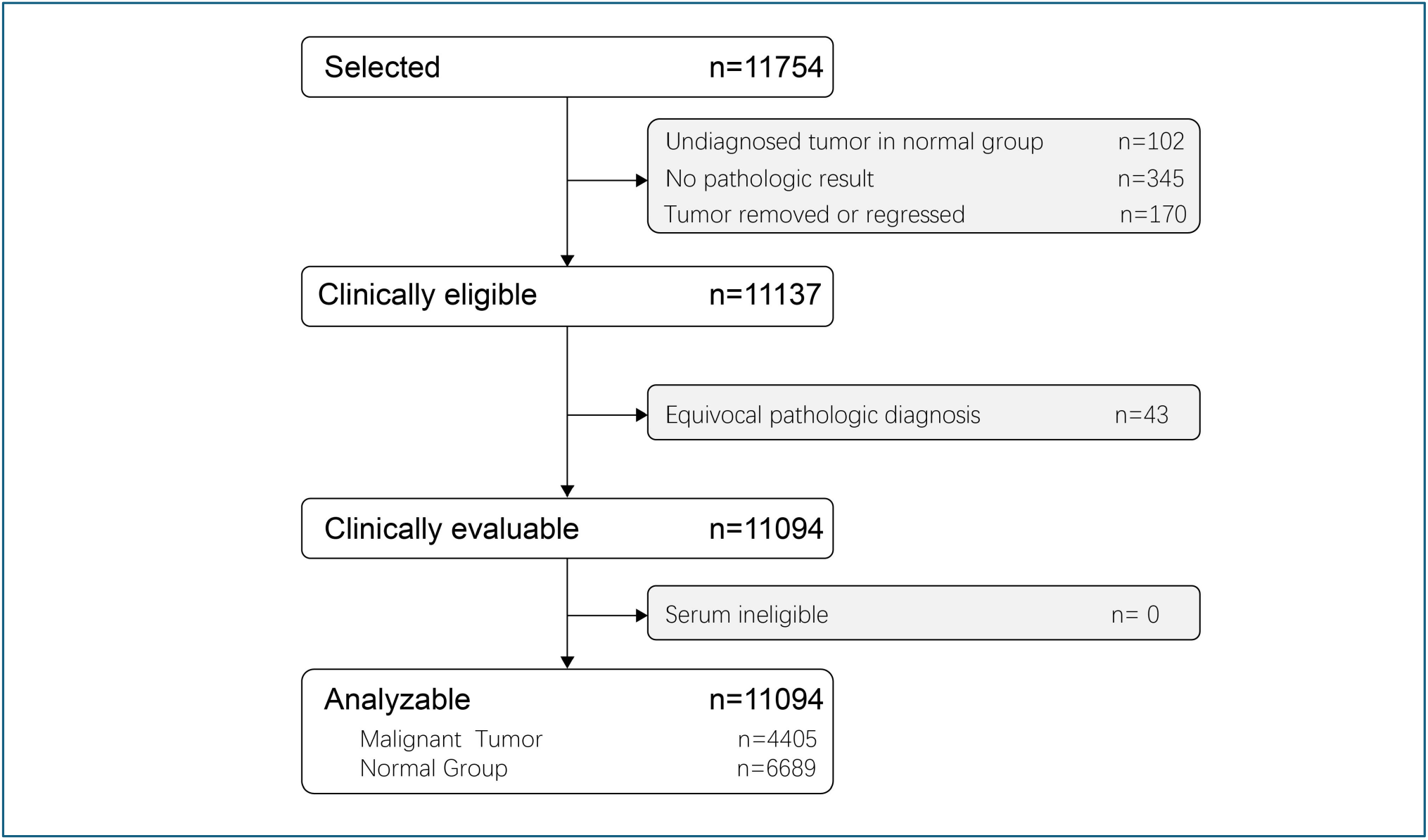
Subject enrollment in this study.

Table 1 lists the demographic and clinical characteristics of the 11094 participants. The two groups of participants were comparable in terms of demographic and clinical characteristics (Table 1). There was an expected difference in age group distribution between the malignant and normal groups (i.e., more cancers than non-cancers in the older age groups). The mean (standard deviation) age was 51.7 (13.44) years and 46.6% (5172/11094) were female (comparable proportions in both groups). 66.9% (2764/4405) of the malignant tumor group were stage I/II.

**Table 1.**
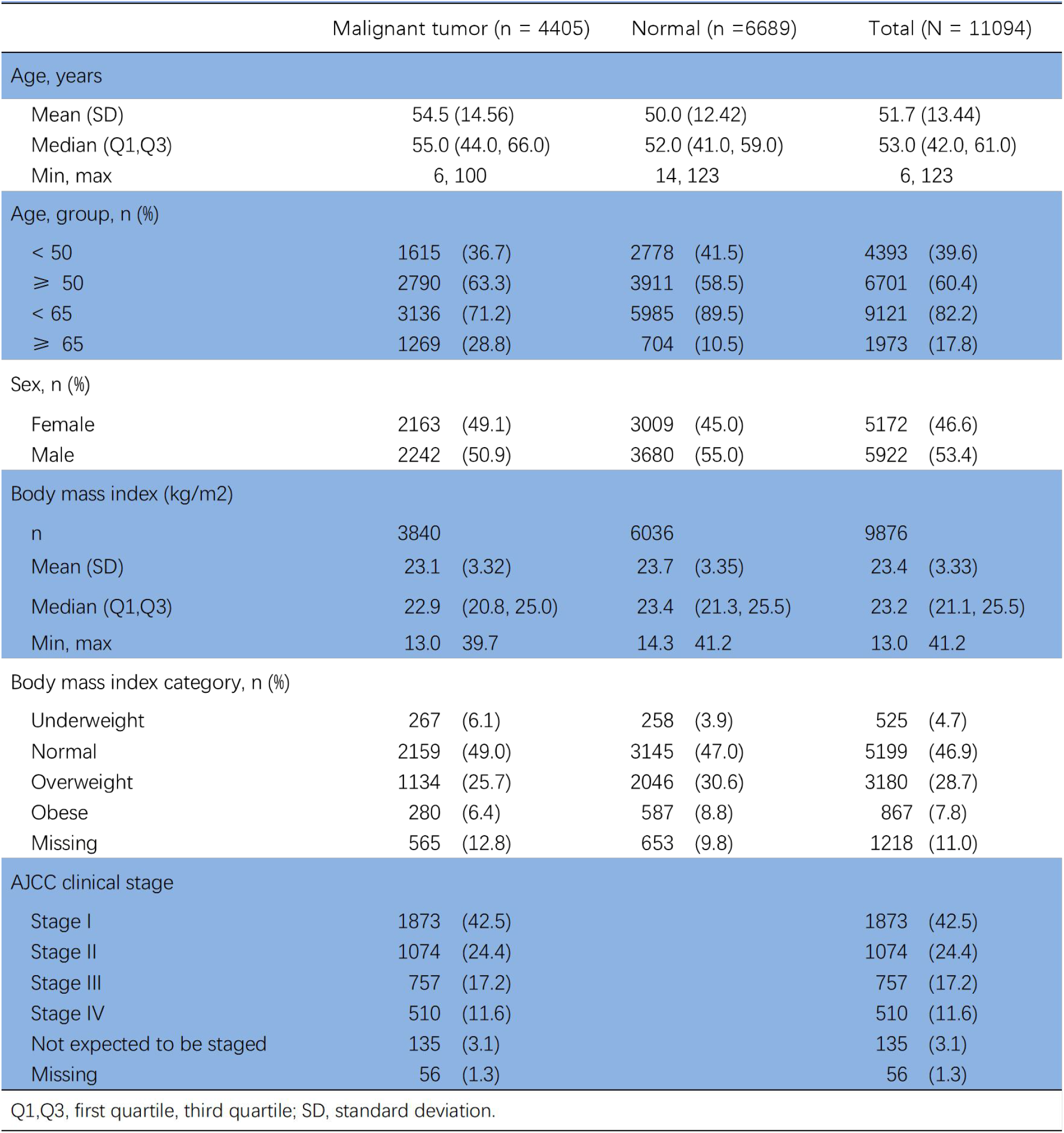
Participants’ demographic and clinical manifestation.

|  | Malignant tumor (n = 4405) | Normal (n = 6689) | Total (N = 11094) |
| --- | --- | --- | --- |
| Age, years |  |  |  |
| Mean (SD) | 54.5 (14.56) | 50.0 (12.42) | 51.7 (13.44) |
| Median (Q1,Q3) | 55.0 (44.0, 66.0) | 52.0 (41.0, 59.0) | 53.0 (42.0, 61.0) |
| Min, max | 6, 100 | 14, 123 | 6, 123 |
| Age, group, n (%) |  |  |  |
| < 50 | 1615 (36.7) | 2778 (41.5) | 4393 (39.6) |
| ≥ 50 | 2790 (63.3) | 3911 (58.5) | 6701 (60.4) |
| < 65 | 3136 (71.2) | 5985 (89.5) | 9121 (82.2) |
| ≥ 65 | 1269 (28.8) | 704 (10.5) | 1973 (17.8) |
| Sex, n (%) |  |  |  |
| Female | 2163 (49.1) | 3009 (45.0) | 5172 (46.6) |
| Male | 2242 (50.9) | 3680 (55.0) | 5922 (53.4) |
| Body mass index (kg/m<sup>2</sup>) |  |  |  |
| n | 3840 | 6036 | 9876 |
| Mean (SD) | 23.1 (3.32) | 23.7 (3.35) | 23.4 (3.33) |
| Median (Q1,Q3) | 22.9 (20.8, 25.0) | 23.4 (21.3, 25.5) | 23.2 (21.1, 25.5) |
| Min, max | 13.0 39.7 | 14.3 41.2 | 13.0 41.2 |
| Body mass index category, n (%) |  |  |  |
| Underweight | 267 (6.1) | 258 (3.9) | 525 (4.7) |
| Normal | 2159 (49.0) | 3145 (47.0) | 5199 (46.9) |
| Overweight | 1134 (25.7) | 2046 (30.6) | 3180 (28.7) |
| Obese | 280 (6.4) | 587 (8.8) | 867 (7.8) |
| Missing | 565 (12.8) | 653 (9.8) | 1218 (11.0) |
| AJCC clinical stage |  |  |  |
| Stage I | 1873 (42.5) |  | 1873 (42.5) |
| Stage II | 1074 (24.4) |  | 1074 (24.4) |
| Stage III | 757 (17.2) |  | 757 (17.2) |
| Stage IV | 510 (11.6) |  | 510 (11.6) |
| Not expected to be staged | 135 (3.1) |  | 135 (3.1) |
| Missing | 56 (1.3) |  | 56 (1.3) |

Q1,Q3, first quartile, third quartile; SD, standard deviation.

The malignant tumor group includes all major solid and hematological malignant tumors: oral cavity, pharynx, larynx, brain, nasopharynx, thyroid, lung, esophagus, stomach, colorectum, breast, liver, gallbladder, pancreas, kidney, urinary tract, ovary, uterus, cervix, prostate, testicle, penis, lymphoma, leukemia1^1,42^. In this study, “others” refers to malignant tumor types other than those listed above, such as adrenal cancer, vulvar cancer, skin cancer, melanoma, metastatic cancer of unknown primary site, etc.

### Performance of YiDiXie™-HS

The overall sensitivity of YiDiXie ™ -HS for different cancer types and stages was 90.1% (95% CI: 89.2% - 90.9%; 3971/4405) and the specificity was 89.7% (95% CI: 89.0% - 90.4%; 6002/6689) (Table 2).

**Table 2.** YiDiXie™-HS test performance between two groups.

|  | Malignant tumor | Normal | Total |
| --- | --- | --- | --- |
|  | 4405 | 6689 | 11094 |
| Test positive | 3971 | 687 | 4658 |
| Test negative | 434 | 6002 | 6436 |

Sensitivity = 3971/4405 90.1% ( 89.2% - 91.0% )
Specificity = 6002/6689 89.7% ( 89.0% - 90.4% )
Two-sided 95% Wilson confidence intervals were calculated.

The sensitivity of YiDiXie™-HS increased with increasing stage in different clinical stages: stage I, 85.6% (83.9% - 87.1%); stage II, 91.4% (89.6% - 93.0%); stage III, 93.9% ( 92.0% - 95.4%); stage IV, 98.4% ( 96.9% - 99.2%) (Table 3). Therefore, YiDiXie™-HS has high sensitivity for all clinical stages.

**Table 3.** Sensitivity of YiDiXie™-HS test by clinical stage.

| Clinical stage | Total N | Test positive | Sensitivity % (95% CI)<sup>a</sup> |
| --- | --- | --- | --- |
| All | 4405 | 3971 | 90.1% ( 89.2% - 91.0% ) |
| I | 1873 | 1603 | 85.6% ( 83.9% - 87.1% ) |
| II | 1074 | 982 | 91.4% ( 89.6% - 93.0% ) |
| III | 757 | 711 | 93.9% ( 92.0% - 95.4% ) |
| IV | 510 | 502 | 98.4% ( 96.9% - 99.2% ) |
| I - II | 2947 | 2585 | 87.7% ( 86.5% - 88.9% ) |
| I - III | 3704 | 3296 | 89.0% ( 87.9% - 90.0% ) |
| I - IV | 4214 | 3798 | 90.1% ( 89.2% - 91.0% ) |
| III - IV | 1267 | 1213 | 95.7% ( 94.5% - 96.7% ) |
| Not expect to be staged | 135 | 123 | 91.1% ( 85.1% - 94.8% ) |
| Missing | 56 | 50 | 89.3% ( 78.5% - 95.0% ) |

CI, confidence interval.
<sup>a</sup>Two-sided 95% Wilson confidence intervals were calculated.

The sensitivity of YiDiXie ™ -HS for different malignant tumor types is shown in Figure 2. The sensitivity of the most of the malignant tumor types range from 82.3% to 96.7%, except for a few cancer types with a small number of cases. Therefore, YiDiXie™-HS has high sensitivity for all cancer types.

**Figure 2.**
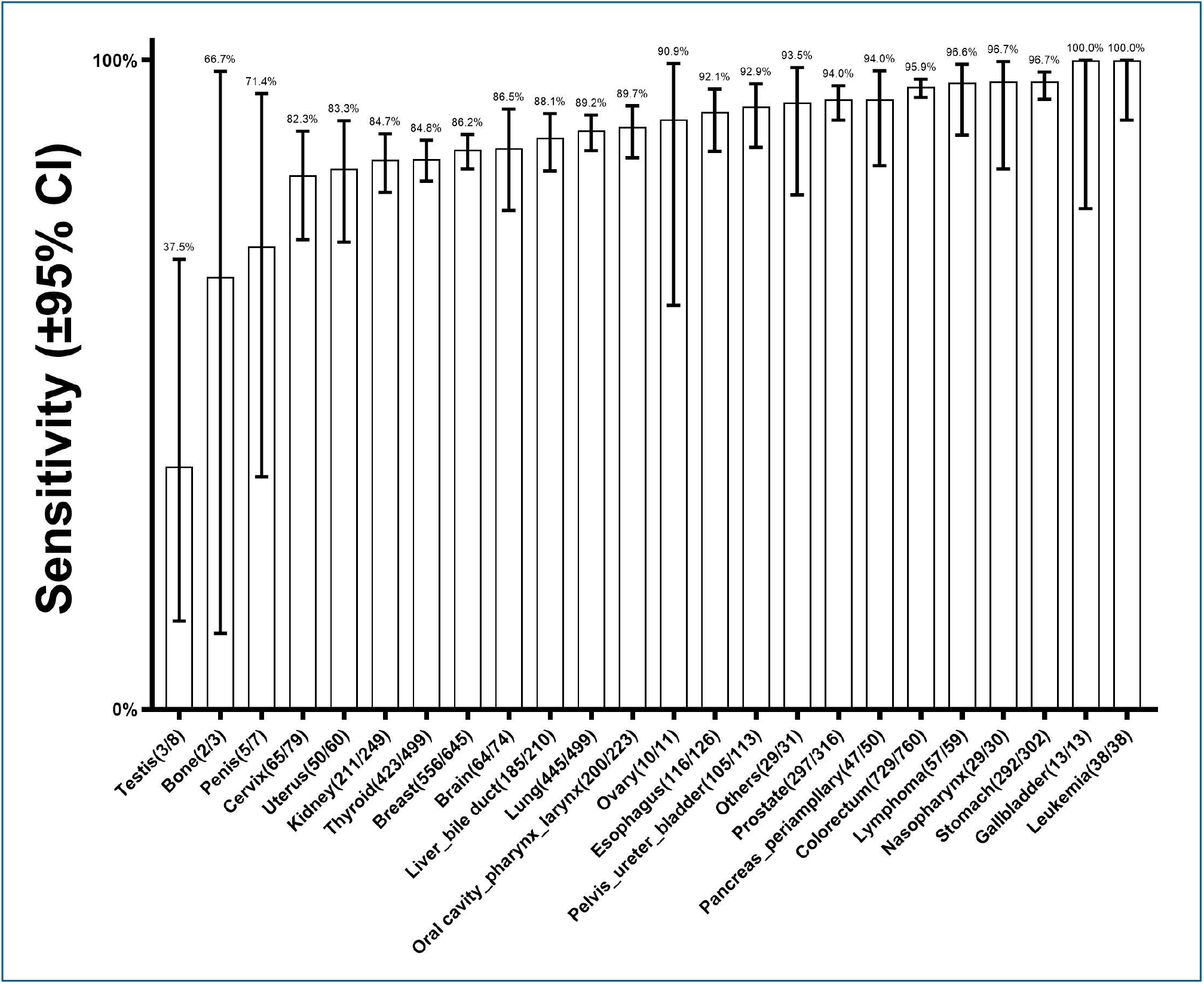
Sensitivity of YiDiXie ™-HS in different malignant tumor types. The horizontal axis shows different malignant tumor types, including all malignant tumor types (including all solid and hematological malignant tumors) covered in “China cancer registry annual report” by the National Cancer Center of China. “Others” are malignant tumor types other than those mentioned above. Bars represent 95% confidence intervals.

Figure 3 reports the sensitivity of YiDiXie™-HS for 16 cancer types with clinical stages and a large number of cases. The results show that YiDiXie ™ -HS has high sensitivity for all stages of these 16 cancer types, except for a few clinical stages with few cases.

**Figure 3.**
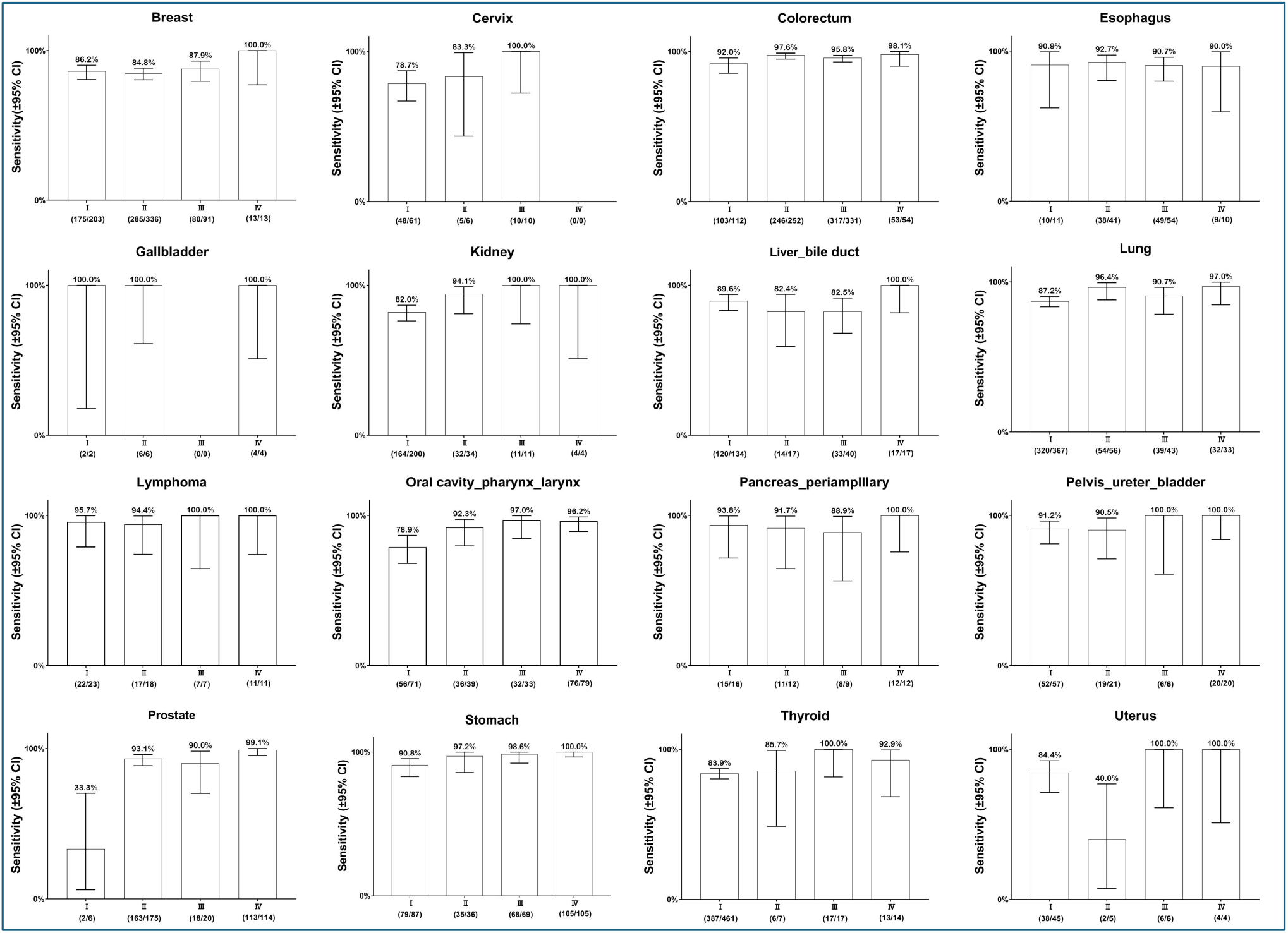
Sensitivity of YiDiXie™-HS in 16 cancer categories with a high number of clinically staged cases. Bars represent 95% confidence intervals.

### Performance of YiDiXie™-SS

The overall sensitivity of YiDiXie ™ -SS for different cancer types and stages was 99.1% (95% CI: 98.8% - 99.3% ; 4365/4405) and its specificity was 65.2% (95% CI: 64.0% - 66.3%; 4358/6689) (Table 4).

**Table 4.** YiDiXie™-SS test performance between two groups.

|  | Malignant tumor | Normal | Total |
| --- | --- | --- | --- |
|  | 4405 | 6689 | 11094 |
| Test positive | 4365 | 2331 | 6696 |
| Test negative | 40 | 4358 | 4398 |

Sensitivity = 4365/4405 99.1% ( 98.8% - 99.3% )
Specificity = 4358/6689 65.2% (64.0% - 66.3% )
Two-sided 95% Wilson confidence intervals were calculated.

The sensitivity of YiDiXie™-SS was equivalent across clinical stages: stage I, 98.6% ( 98.0% - 99.1% ); stage II, 99.5% ( 98.9% - 99.8% ); stage III, 99.5% (98.6% - 99.8%); stage IV, 99.8% ( 98.9% - 99.8% ) (Table 5). Therefore, YiDiXie ™-SS has a very high sensitivity for all clinical stages.

**Table 5.** Sensitivity of YiDiXie™-SS test by clinical stage.

| Clinical stage | Total N | Test positive | Sensitivity % (95% CI)<sup>a</sup> |
| --- | --- | --- | --- |
| All | 4405 | 4365 | 99.1% ( 98.8% - 99.3% ) |
| I | 1873 | 1847 | 98.6% ( 98.0% - 99.1% ) |
| II | 1074 | 1069 | 99.5% ( 98.9% - 99.8% ) |
| III | 757 | 753 | 99.5% ( 98.6% - 99.8% ) |
| IV | 510 | 509 | 99.8% ( 98.9% - 100.0% ) |
| I - II | 2947 | 2916 | 98.9% ( 98.5% - 99.3% ) |
| I - III | 3704 | 3669 | 99.1% ( 98.7% - 99.3% ) |
| I - IV | 4214 | 4178 | 99.1% ( 98.8% - 99.4% ) |
| III - IV | 1267 | 1262 | 99.6% ( 99.1% - 99.8% ) |
| Not expect to be staged | 135 | 133 | 98.5% ( 94.8% - 99.7% ) |
| Missing | 56 | 54 | 96.4% ( 87.9% - 99.4% ) |

CI, confidence interval.
<sup>a</sup> Two-sided 95% Wilson confidence intervals were calculated.

The sensitivity of YiDiXie ™ -SS for different malignant tumor types is shown in Figure 4. The results showed that the sensitivity for various cancer types ranged from 97.3% to 100%. Therefore, YiDiXie™-SS has very high sensitivity in all cancer types.

**Figure 4.**
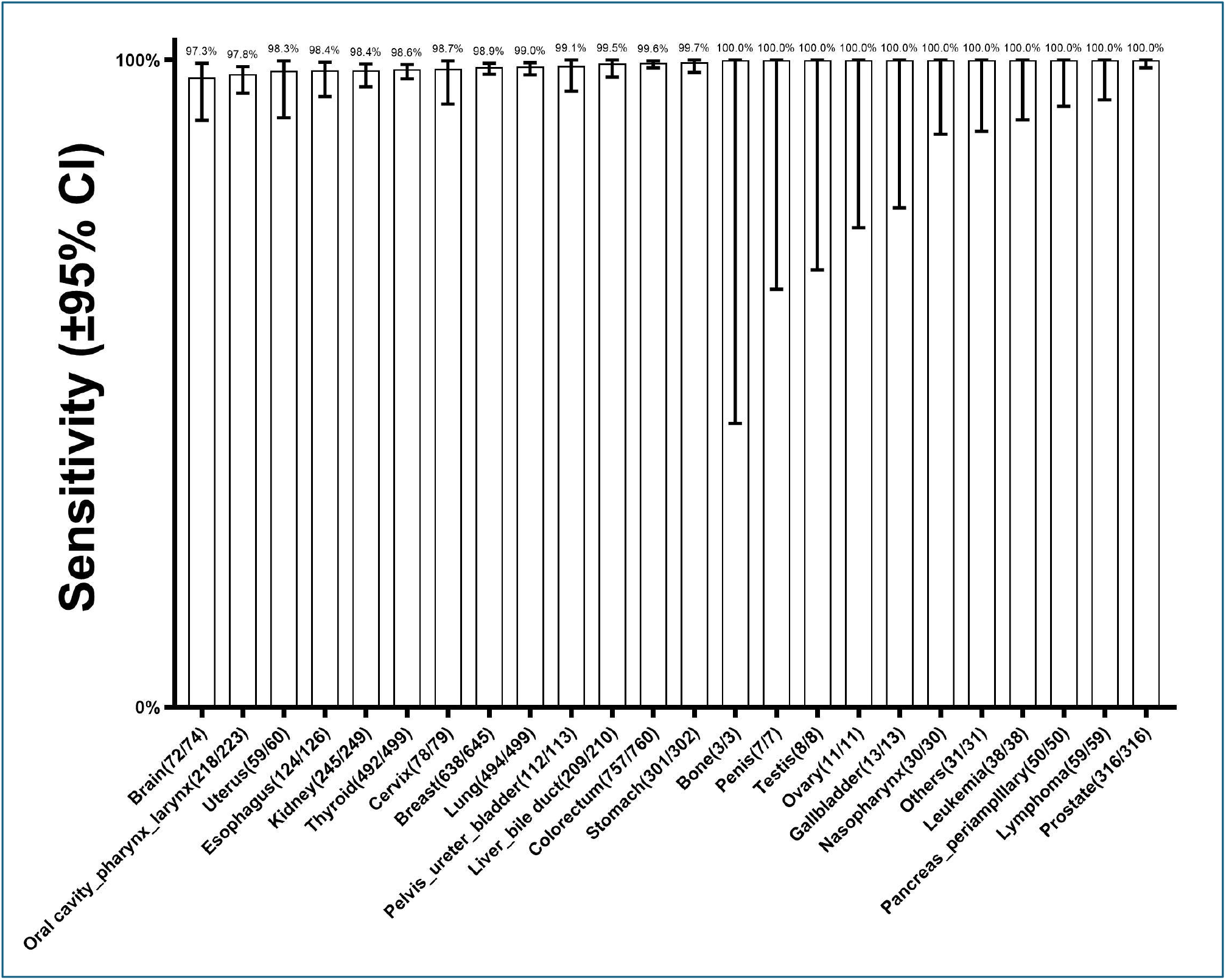
Sensitivity of YiDiXie ™ -SS in different malignant tumor types. The horizontal axis shows different malignant tumor types, including all malignant tumor types (including all solid and hematological malignant tumors) covered in “China cancer registry annual report” by the National Cancer Center of China. “Others” are malignant tumor types other than those mentioned above. Bars represent 95% confidence intervals.

Figure 5 reports the sensitivity of YiDiXie™-SS for 16 cancer types with clinical stages and a large number of cases. The results show that YiDiXie ™ -SS has very high sensitivity in all stages of these 16 cancers.

**Figure 5.**
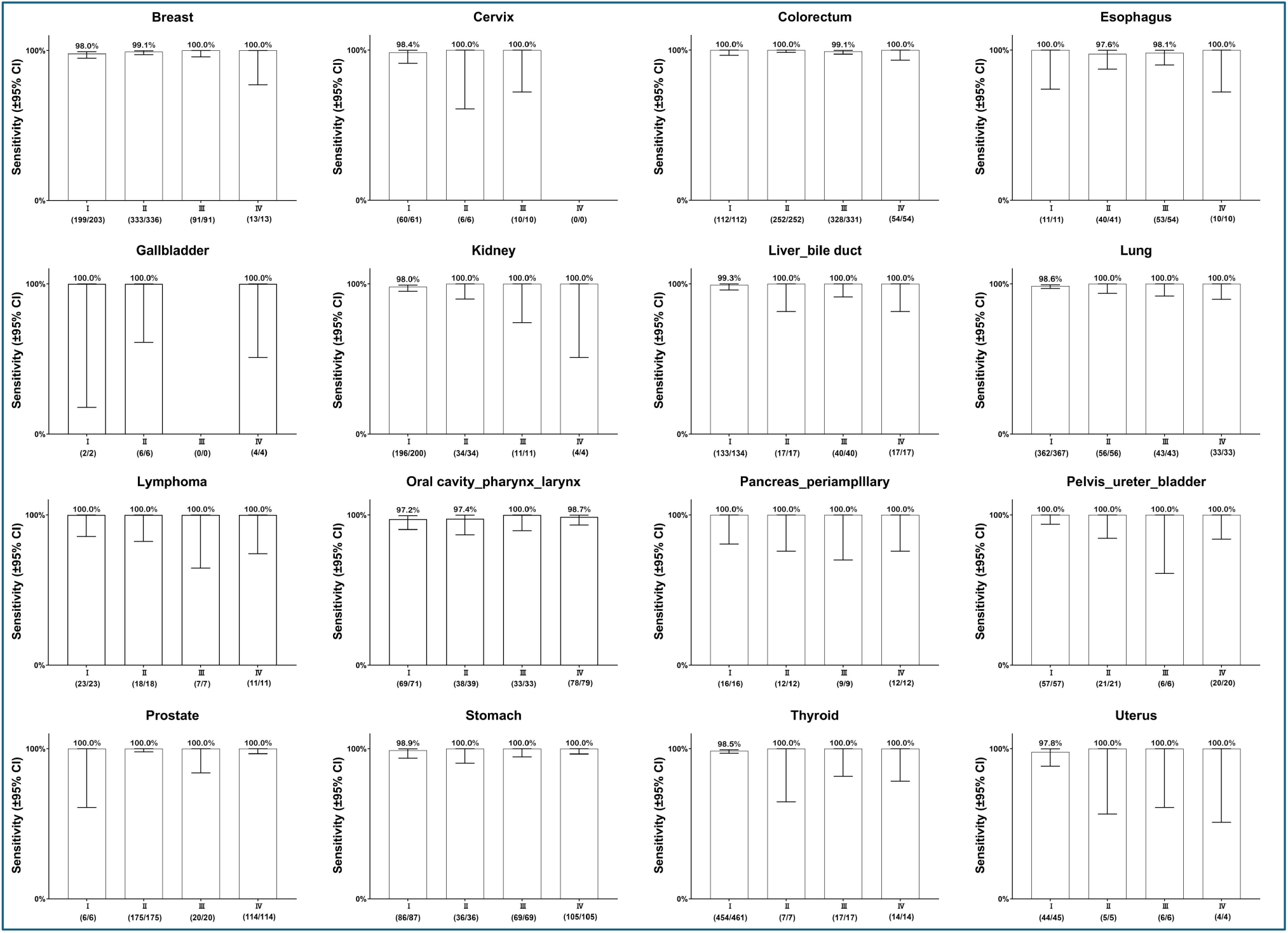
Sensitivity of YiDiXie™-SS in 16 cancer categories with a high number of clinically staged cases. Bars represent 95% confidence intervals.

## DISCUSSION

### MCED model is expected to solve the challenge of “Universal Cancer Screening”

Currently, the published MCED tests are Galleri™ test^43^, CancerSEEk ™ test^44^, DEEPGEN ™ test^45^, PanSEER test^46^. cfMeDIP-Seq test^47^, IvyGene^®^ test^48^. The most representative of these are CancerSEEk™ test^44 49^by Thrive, Inc., developed based on cfDNA and protein-based tumor markers, and Galleri ™ test ^43^ by Grail, Inc., developed based on cfDNA.

It is the emerging MCED test that makes the MCED model a reality. Compared to the SCED model, the MCED model has the following advantages: (1) Having a broader cancer screening spectrum and aggregated prevalence^20^. (2) Significant improving mental health and decrease subsequent medical costs with reduction in false-positive results^20^. (3) Having a higher early cancer detection rate^50,51^. (4) Significantly reduction cancer-specific mortality^10,51^. (5) Significantly improving the prognosis of cancer patients^51^ and reducing the cost of cancer treatment^52^. (6) More acceptable to the public^53^.

In fact, the MCED model based on the MCED test may be the only cost-effective screening model for lower-prevalence cancer types^9^ and is expected to solve the world’s “Universal cancer screening” problem.

### The nature of the MCED model

The MCED test treats multiple cancer types as a single disease type, “cancer or malignant tumor”, and converts “multiple screenings for multiple cancers” into “a single screening for multiple cancers”. Compared to the SCED model, which can only screen for a single cancer with conventional imaging (e.g., CT scan of the lungs) or testing products (e.g., blood TPSA), the MCED model can screen for multiple cancers throughout the body with a single MCED test.

All MCED tests screen for cancer via the MCED model. The MCED model is essentially an MCED test that serves as the “primary screening”, which is followed by the “secondary screening”. In short, the MCED test as the “primary screening” unifies multiple cancer types into a single disease type of “cancer or malignant tumor”; The MCED test only determines “whether it is cancer”, not “where it is located”. A “negative” result of the MCED test ends the screening, and a “positive” result is followed by “secondary screening” to determine “where the cancer is located.”

The “primary screening” of the MCED model is an MCED test, such as the CancerSEEK ™ test, the Galleri ™ test, the “YiDiXie ™ test”, etc. The “secondary screening” for the MCED model is a test or group of tests. The “secondary screening” for the CancerSEEK ™ test is PET-CT^44^. The “secondary screening” for Galleri ™ test is a self-developed localization system ^43^, The “secondary screening” of the “YiDiXie™ test” is a package of routine medical checkups for comprehensive cancer screening (including physical examination, ultrasound, CT, MRI, blood TPSA, etc.).

### The rationale behind the development of the two MCED tests

An MCED test is ideal if it has both extreme sensitivity and extreme specificity. However, sensitivity and specificity of the same test are a contradiction in terms. Therefore, developers often must balance the pros and cons of prioritizing sensitivity or specificity.

Both the sensitivity and specificity of an MCED test are critical. On the one hand, the sensitivity of an MCED test is essential; lower sensitivity means a higher false-negative rate. Since screening ends with a negative MCED test result, a higher false-negative rate means that more cases are missed. This will most likely lead to developing advanced cancer and result in a series of adverse consequences such as poor prognosis and significantly increased social and patient economic burden.

On the other hand, the specificity of an MCED test is very important, and a lower specificity means a higher rate of false positives. Since “secondary screening” is required when the result of the MCED test is positive, a higher false-positive rate undoubtedly increases the cost of “secondary screening” significantly, which obviously increases the economic burden on society and the examinees.

Thus, the balance between the sensitivity and specificity of an MCED test is essentially a balance between “fewer cases missed” and “lower costs of secondary screening”.

In general, payers such as governments, insurance companies, and charitable organizations choose cancer screening products with better cost-benefit analyses based on the perspectives of health economics. These payers prefer products that combine the advantages of “fewer cases missed” and “lower costs of secondary screening”.

For this reason, YiDiXie™-HS is optimized for both sensitivity and specificity in the development process. As shown in Table 2, the overall sensitivity of YiDiXie™-HS across cancer types and stages was 90.1% (95% CI: 89.2% - 90.9%; 3971/4405), while the specificity was 89.7% (95% CI: 89.0% - 90.4%; 6002/6689). Thus, YiDiXie™-HS optimally combines sensitivity and specificity.

Accordingly, YiDiXie™-HS has the advantages of both “fewer cases missed” and “lower costs of secondary screening”, making it suitable for payers such as governments, commercial insurers, and charitable organizations that are focused on cost-performance analyses.

Nevertheless, the cost of “secondary screening” is not sensitive to non-paying recipients with high screening costs or paying recipients in good financial circumstances. Because of the insensitivity to the cost of “secondary screening”, these subjects prefer the MCED test to detect as many cases as possible and to avoid missing cases if possible. In other words, for these subjects, “fewer cases missed” is much more crucial than “lower costs of secondary screening”. Therefore, these patients need the MCED test, which has a extremely high sensitivity and a relatively low specificity.

Thus, YiDiXie™-SS was developed with the aim of “prioritizing the sensitivity”, which is extremely sensitive with relatively low specificity. YiDiXie™-SS dramatically increases the number of miRNA markers to achieve extremely high sensitivity to all cancer types. As shown in Table 4, the overall sensitivity of YiDiXie™-SS for different cancer types and stages was 99.1% (95% CI: 98.8% - 99.3%; 4365/4405); the specificity was 65.2% (95% CI: 64.0% - 66.3%; 4358/6689). 3895/6005). YiDiXie™-SS well fulfills the development intent.

In brief, YiDiXie ™ -HS has the advantages of both “fewer cases missed” and “lower costs of secondary screening”, making it suitable for payers such as governments, commercial insurers, and charitable organizations that are focused on cost-performance analyses. Accordingly, YiDiXie ™ -SS is ideal for cost-insensitive subjects owing to the “minimal missed cases” but “higher costs of secondary screening”.

### YiDiXie™-HS and YiDiXie™-SS can replace existing cancer screening tests and are expected to solve the world problem of “Universal cancer screening”

Firstly, YiDiXie™-HS and YiDiXie™-SS test can replace existing cancer screening tests. Since patients with early-stage cancers missed during cancer screening are very likely to develop advanced cancers, the only alternative to existing cancer screening tests is the MCED test, which is highly sensitive to all clinical stages of all cancer types, including early-stage cancers.

As the results show, The total sensitivity of YiDiXie ™ -HS test for the malignant tumor group was 90.1% (95% CI: 89.2% - 90.9%; 3971/4405) (Table 2), and the sensitivity was high for all clinical stages: stage I, 85.6% (83.9% - 87.1%); stage II, 91.4% (89.6% - 93.0%); stage III, 93.9% (92.0% - 95.4%); and stage IV, 98.4% (96.9% - 99.2%) (Table 3). While YiDiXie™-SS had an overall sensitivity of 99.1% (95% CI: 98.8% - 99.3% ; 4365/4405) for the malignant tumor group (Table 4), with high sensitivity for all clinical stages: stage I, 98.6% (98.0% - 99.1%); stage II, 99.5% (98.9% - 99.8%); stage III, 99.5% (98.6% - 99.8%); stage IV, 99.8% (98.9% - 99.8%) (Table 5); Therefore, YiDiXie™ -HS and YiDiXie ™ -SS can replace existing cancer screening tests due to their high sensitivity to all clinical stages of all cancer types, including early-stage cancers.

Secondly, the “YiDiXie™ test” requires only a tiny amount of blood, allowing for cancer screening without having to leave one’s home. Only 20 microliters of serum is required to complete a “YiDiXie™ test”, which is equivalent to the volume of 1 drop of whole blood (1 drop of whole blood is about 50 microliters, which produces 20-25 microliters of serum). Considering the pre-test sample quality assessment and 2-3 repetitions of the test, 0.2 ml of whole blood is sufficient to complete the “YiDiXie™ test”. A normal subject can collect 0.2 ml of finger blood at home using a finger blood collection needle without venous blood collection by medical staff. Therefore, the “YiDiXie™ test” allows for cancer screening without having to leave one’s home.

Finally, the “YiDiXie ™ test” has a nearly unlimited cancer screening capacity, allowing for “Universal cancer screening” once a year. The traditional SCED model, whose screening capacity is directly dependent on the number of doctors and equipment, makes it almost impossible to realize “Universal cancer screening” once a year for most cancer types. Figure 6 shows the basic flowchart of the “MCED model of YiDiXie™”, which shows that the “YiDiXie ™ test” does not require not only doctors and medical equipment, but also medical personnel to collect blood. The patient only needs to place an order online, collect 0.2 ml of finger blood at home, and express it to the laboratory to complete the “YiDiXie™ test”. Thus, the “YiDiXie ™ test” enables “Universal cancer screening” once a year.

**Figure 6.**
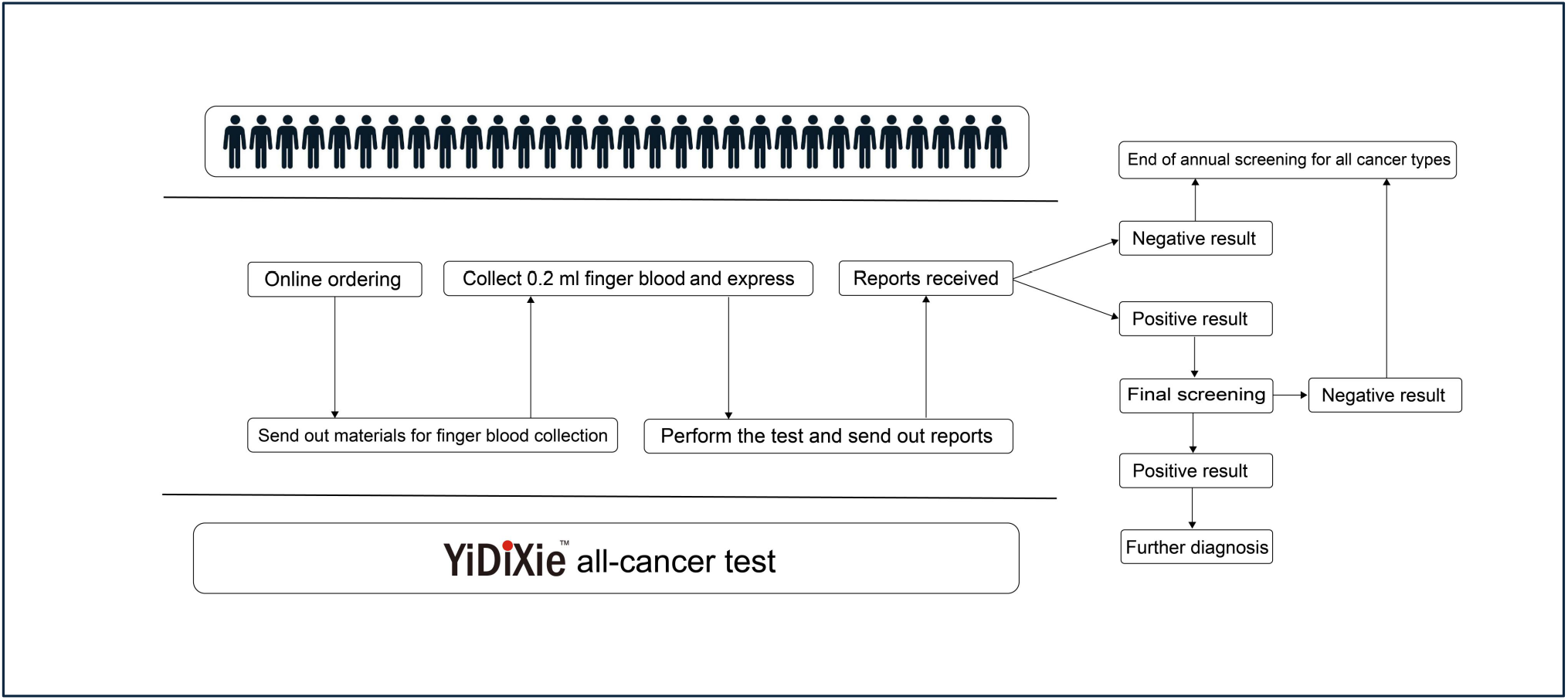
Basic flowchart of the “YiDiXie™ test”.

Consequently, YiDiXie™-HS and YiDiXie™-SS can replace existing cancer screening tests and are expected to solve the world problem of “Universal cancer screening”.

### Limitations of the study

First, the normal control group were the medical examiners who underwent health checkups. The normal control group subjects were not followed up for more than 1 year, so there must be some malignant tumors hidden among them. As a result, the false-positive rate was higher than the actual situation, resulting in a certain bias.

Second, this study was a case-control study, not a cross-sectional study of the normal population. Therefore, this study cannot demonstrate the positive and negative predictive values of the “YiDiXie ™ test” in screening for all malignant tumor types in normal populations.

Final, this study was a single-center, observational study, which could be subject to some bias. In future, multi-center, randomized controlled trials are needed to further evaluate the performance of the “YiDiXie™ test” in screening for all malignant tumor types in normal populations.

## CONCLUSION

YiDiXie™-HS has a high sensitivity in all clinical stages of all cancer types. YiDiXie ™ -SS has an extremely high sensitivity in all clinical stages of all cancer types. YiDiXie ™ -HS and YiDiXie ™ -SS can replace existing cancer screening tests and are expected to solve the world problem of “Universal cancer screening”.

## Data Availability

All data produced in the present study are contained in the manuscript.

